# Analgesic quality and safety of selective supraclavicular nerves and C5 root blockade in clavicle surgery - A randomized controlled trial

**DOI:** 10.1101/2025.09.22.25336305

**Authors:** Martin Julian Schaefer, Martin Zoremba

**Author notes:** **Correspondence to:** Martin Julian Schaefer, Department of Anesthesiology and Intensive Care Medicine, Klinikum Siegen, Academic Teaching Hospital of the University of Marburg, Weidenauerstrasse 76, 57076 Siegen, Germany. **Trial registration:** German Clinical Trial Registry DRKS ID: DRKS00017286, https://drks.de/search/en/trial/DRKS00017286/entails, date of registration May 20, 2019). **Ethics committee approval:** Ethical Commission Westfalen-Lippe, Gartenstrasse 210-214, Münster/Germany, protocol number 2018-645-f-S, date of approval: March 14, 2019, chairperson Univ.-Prof. Dr. med. W.E. Berdel.

## Abstract

**Background:** Clavicle fractures and acromioclavicular (AC) joint dislocations are common in athletic adults. Although peripheral nerve blocks are widely used in upper extremity surgery, their role in clavicle surgery remains debated due to the complex innervation. This study assessed whether selective blockade of the supraclavicular nerves and C5 root is a suitable technique for improving postoperative pain management.

**Methods:** In this randomized trial (German Clinical Trial Registry DRKS ID: DRKS00017286), 56 patients undergoing clavicle or AC joint surgery were allocated to three groups. The first group received general anesthesia with a combined nerve block of the supraclavicular nerves and C5 root (NSCL+C5), the second received general anesthesia only with C5 root block (C5), and the third had general anesthesia with systemic analgesia (control group). Pain levels (NRS) and piritramide consumption were the primary endpoints, while secondary endpoints included diaphragmatic excursion and oxygen saturation to assess phrenic nerve palsy.

**Results:** Patients in the NSCL+C5 group reported no pain during the first postoperative hour (NRS t0-t1=0), significantly less than the C5 (NRS t0=1.9 ± 1.4, *P*=0.001) and control group (NRS t0=4.5 ± 2.2, *P*<0.001). No additional opioid was required, which was significantly lower compared with 4.4 mg ± 5.3 (*P*=0.01) in the C5 group and 12.6 mg ± 5.3 (*P*<0.001) in the control group. Phrenic nerve palsy was more frequent in the NSCL+C5 group, without affecting oxygen saturation.

**Conclusion:** Combined blockade of the supraclavicular nerves and C5 root significantly reduces postoperative pain and opioid consumption in clavicle surgery. Despite a higher rate of phrenic palsy, respiratory function remained unaffected.

## Introduction

Clavicle fractures and acromioclavicular (AC) joint dislocations are common injuries in active adults and account for a significant proportion of shoulder girdle trauma.^1,2^ Surgical treatment of these injuries is typically performed under general anesthesia with systemic opioid-based analgesia. However, the routine use of peripheral nerve blocks in this context remains controversial due to the complex innervation of the clavicle, which involves both the cervical and brachial plexuses.^3-6^ This complexity has led to ongoing debate regarding the optimal regional anesthesia technique, and establishing a standardized approach for peri- and postoperative pain management in clavicle fractures and AC joint dislocations remains challenging.

Considering the proven benefits of regional anesthesia—such as superior analgesia, reduced opioid consumption, and enhanced postoperative recovery^7,8^—optimizing its application for clavicle injuries is both necessary and clinically valuable. Nonetheless, brachial and cervical plexus blocks, particularly the interscalene approach, are associated with a high incidence of transient hemidiaphragmatic paralysis due to the proximity of the phrenic nerve.^9-11^ While this is usually well tolerated in healthy individuals, it may pose a risk to patients with pre-existing pulmonary conditions.^9-11^

Based on current anatomical and clinical knowledge, we hypothesize that a combined regional anesthesia technique targeting the supraclavicular nerves and the C5 nerve root will result in significantly lower postoperative pain scores (NRS) and reduced opioid consumption compared with standard systemic analgesia. Additionally, this study investigates whether this technique— despite a potentially higher incidence of phrenic nerve palsy—has any clinically relevant impact on postoperative oxygen saturation.

## Methods

### Ethics approval and randomization

Following approval by the Ethics Committee (Ethical Commission Westfalen-Lippe, Gartenstrasse 210-214, Münster/Germany, protocol number 2018-645-f-S, date of approval: March 14, 2019) and registration in the German Clinical Trial Register (DRKS ID: DRKS00017286, date of registration May 20, 2019), this randomized controlled trial was conducted in accordance with the latest version of the Declaration of Helsinki at the Department of Anesthesiology of the Klinikum Siegen, Academic Teaching Hospital of the University of Marburg, Germany. A total of 60 patients were enrolled between May 27, 2019 and November 30, 2020. Eligibility criteria for study participation included patients (ASA classification I to III) scheduled for elective surgical treatment of a clavicle fracture or AC joint dislocation. Exclusion criteria were age under 18 years, inability to provide consent, pregnancy, pre-existing phrenic nerve palsy, and known contraindications to regional anesthesia. After providing both verbal and written informed consent, patients were randomly assigned to one of three study groups in an open-label manner. The first group (NSCL+C5) underwent general anesthesia in combination with a selective regional blockade of both the supraclavicular nerves and the C5 nerve root. The second group (C5) received general anesthesia supplemented by a selective C5 root block alone. The third group (control) was treated with general anesthesia and systemic opioid-based analgesia.

### Study design and practical information

Patients were recruited within 24 hours before surgery during the anesthesiological premedication visit or on the surgical wards. During this visit, patients underwent a routine pre-operative assessment, which included medical history, physical examination, and detailed study information. A layperson-friendly informational brochure was provided. Patients were enrolled only after giving both written and oral informed consent and fulfilling all inclusion and exclusion criteria. Demographic data such as age, sex, weight, height, and BMI were recorded. Randomization was performed in the holding area using sealed envelopes by an independent anesthesia nurse.

Before anesthesia induction, baseline measurements were taken, including standard monitoring, pain assessment, and diaphragmatic excursion via ultrasound. Patients in the intervention groups received ultrasound-guided regional anesthesia (supraclavicular nerves and/or C5 root block) prior to general anesthesia, while the control group received no regional anesthesia. General anesthesia was induced with propofol (1.5–2.5 mg kg^-1^), fentanyl (2–5 µg kg^-1^), and rocuronium (0.3 mg kg^-1^) for intubation. Anesthesia maintenance included remifentanil (0.2 µg kg^-1^ min^-1^, continuous infusion) and desflurane (3–4 vol%). To prevent postoperative nausea and vomiting (PONV), all patients received dexamethasone (4 mg) and granisetron (1 mg). In the control group, metamizole (30 mg kg^-1^) was administered intraoperatively for baseline analgesia. After emergence from anesthesia, patients were transferred to the post-anesthesia care unit (PACU), where the first postoperative assessment (t0) was performed. Further assessments followed after 30 minutes (t0.5) and 60 minutes (t1). Pain management targeted an NRS≤4 and was achieved via piritramide (3.75 mg bolus doses as needed). A final assessment 24 hours postoperatively (t24) included pain score (NRS), oxygen saturation (pulse oximetry), and diaphragmatic excursion (ultrasound).

### Regional anesthesia technique

All regional anesthesia procedures were performed using an ultrasound-guided in-plane technique with a linear probe (Sonosite X-Porte HFL 50xp, 15-6 MHz, FUJIFILM Sonosite Inc., Bothell, WA, USA) and a 50 mm needle (Ultraplex® 360, B. Braun Melsungen AG, Melsungen, Germany) through a single skin puncture (Figure 1). The supraclavicular nerves were identified anterior to the prevertebral fascia and middle scalene muscle, embedded in the cervical fascia. For effective blockade, 5 ml of local anesthetic (2.5 ml ropivacaine 0.75% + 2.5 ml prilocaine 1%) was injected to ensure fascial spread. The needle was then advanced under ultrasound guidance to the C5 nerve root, where an additional 5 ml of the same anesthetic mixture was administered. Patients in the NSCL+C5 group received a total of 10 ml (37.5 mg ropivacaine + 50 mg prilocaine), while those in the C5 group received 5 ml (18.75 mg ropivacaine + 25 mg prilocaine).

**Figure 1.**
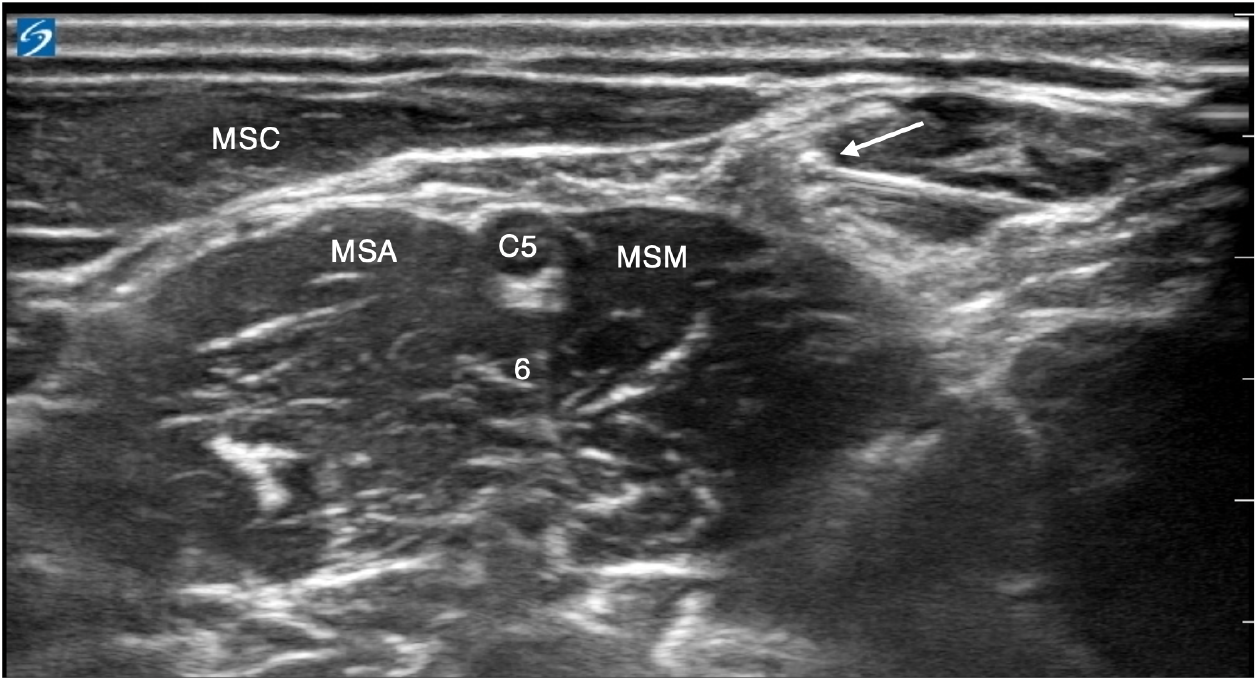
Selective blockade of the supraclavicular nerves and the C5 root. Arrow= needle positioning at the level of the supraclavicular nerves, C5= C5 nerve root, 6= C6 nerve root, MSC= sternocleidomastoid muscle, MSA= anterior scalene muscle, MSM= middle scalene muscle

### Primary endpoints: Postoperative pain levels and opioid consumption

Postoperative pain was assessed using the Numeric Rating Scale (NRS) at four time points: t0 (immediately postoperative), t0.5 (30 minutes), t1 (60 minutes), and t24 (24 hours postoperative). Patients rated pain from 0 (no pain) to 10 (worst imaginable pain). An NRS>3 prompted the administration of additional analgesia. Piritramide was used for pain control with an initial bolus of 3.75 mg administered for NRS>3, followed by further doses titrated as needed. The cumulative 24-hour dose was recorded as an objective measure of analgesic requirements.

### Secondary endpoints: Diaphragmatic excursion and oxygen saturation

To assess phrenic nerve function, diaphragmatic excursion was measured using M-mode ultrasound with a 1–5 MHz curvilinear abdominal probe (FUJIFILM Sonosite Inc., Bothell, WA, USA). Excursion during quiet breathing was recorded, with probe position marked on the patient’s skin for consistency. Measurements were taken pre- and postoperatively (t0–t24). Oxygen saturation was monitored via pulse oximetry and recorded at specified intervals under room air during quiet, supine breathing. All postoperative values were expressed relative to baseline.

### Statistics

The sample size calculation (Number Cruncher Statistical Systems, PASS 2002 Software, Kaysville, UT, USA) was based on an assumed standard deviation of σ=1.5, an alpha error probability of 5% (α=0.05), and a power of >80% (β error<0.2). This resulted in a required sample size of 17 patients per study group. Assuming a strong effect size (f=0.45), with a statistical power of 0.80 and an alpha error probability of 0.05 (α=0.05), the recommended sample size was 17 patients per group, as calculated using G*Power software (Version 3.1; Heinrich Heine University Düsseldorf, Germany). To account for potential patient exclusions and enhance data stability, the sample size was increased to 20 patients per study group before study initiation. Statistical analysis was performed using SPSS software (IBM SPSS Statistics for Windows, Version 26.0; IBM Corp., Armonk, NY, USA). Group comparisons were performed using one-way analysis of variance (ANOVA), followed by post hoc testing with Bonferroni correction to adjust for multiple comparisons and control for the family-wise error rate. The formulated hypotheses regarding the primary and secondary endpoints were tested at a significance level of *P*≤0.05. The collected study data were transcribed from handwritten case report forms into a spreadsheet using Microsoft Excel (version 16.16.18; Microsoft Corp., Redmond, WA, USA).

## Results

A total of 60 patients were enrolled between May 27, 2019 and November 30, 2020. In two cases, the planned surgery was cancelled in favor of conservative treatment. Two additional patients, despite initially providing consent, declined the regional anesthesia procedure at short notice and were therefore excluded. Ultimately, 56 patients were randomized into three study groups. No further exclusions occurred during data collection or statistical analysis, as shown in the flow diagram (Figure 2). Baseline characteristics are presented in Table 1.

**Table 1.** Characteristics of the patients at baseline.

|  | NSCL+C5<br>(n=19) | C5<br>(n=18) | control<br>(n=19) |
| --- | --- | --- | --- |
| Age (years) | 38 ± 13 | 44 ± 16 | 33 ± 15 |
| Height (cm) | 179 ± 8 | 176 ± 10 | 178 ± 9 |
| Weight (kg) | 84 ± 14 | 75 ± 13 | 80 ± 15 |
| BMI (kg.m<sup>-2</sup>) | 26 ± 4 | 24 ± 3 | 25 ± 4 |
| Sex, male/female (n) | 15/4 | 12/6 | 15/4 |
| ASA status 1/2/3 (n) | 8/11/0 | 9/9/0 | 11/8/0 |
| NRS pre-op | 1.2 ± 1.3; (1) | 1.2 ± 1.2; (1) | 1.4 ± 1.8; (1) |
| DE pre-op (cm) | 2.3 ± 0,4 | 2.1 ± 0,2 | 2.4 ± 0.4 |
| SpO<sub>2</sub> pre-op (%) | 98.5 ± 1,5 | 98.5 ± 1,6 | 98.8 ± 1.0 |
| Surgery time (min) | 56 ± 27 | 67 ± 30 | 64 ± 23 |
| Clavicle fracture/AC joint dislocation (n) | 12/7 | 12/6 | 17/2 |

*Values are expressed as mean ± SD; (median) or absolute number (n); NSCL+C5=combined supraclavicular nerves and C5 root blockade, C5=C5 root blockade, control=systemic analgesia group, BMI=body mass index; AC=acromioclavicular, NRS=numeric rating scale, DE=diaphragmatic excursion, pre-op=pre-operative*

**Figure 2.**
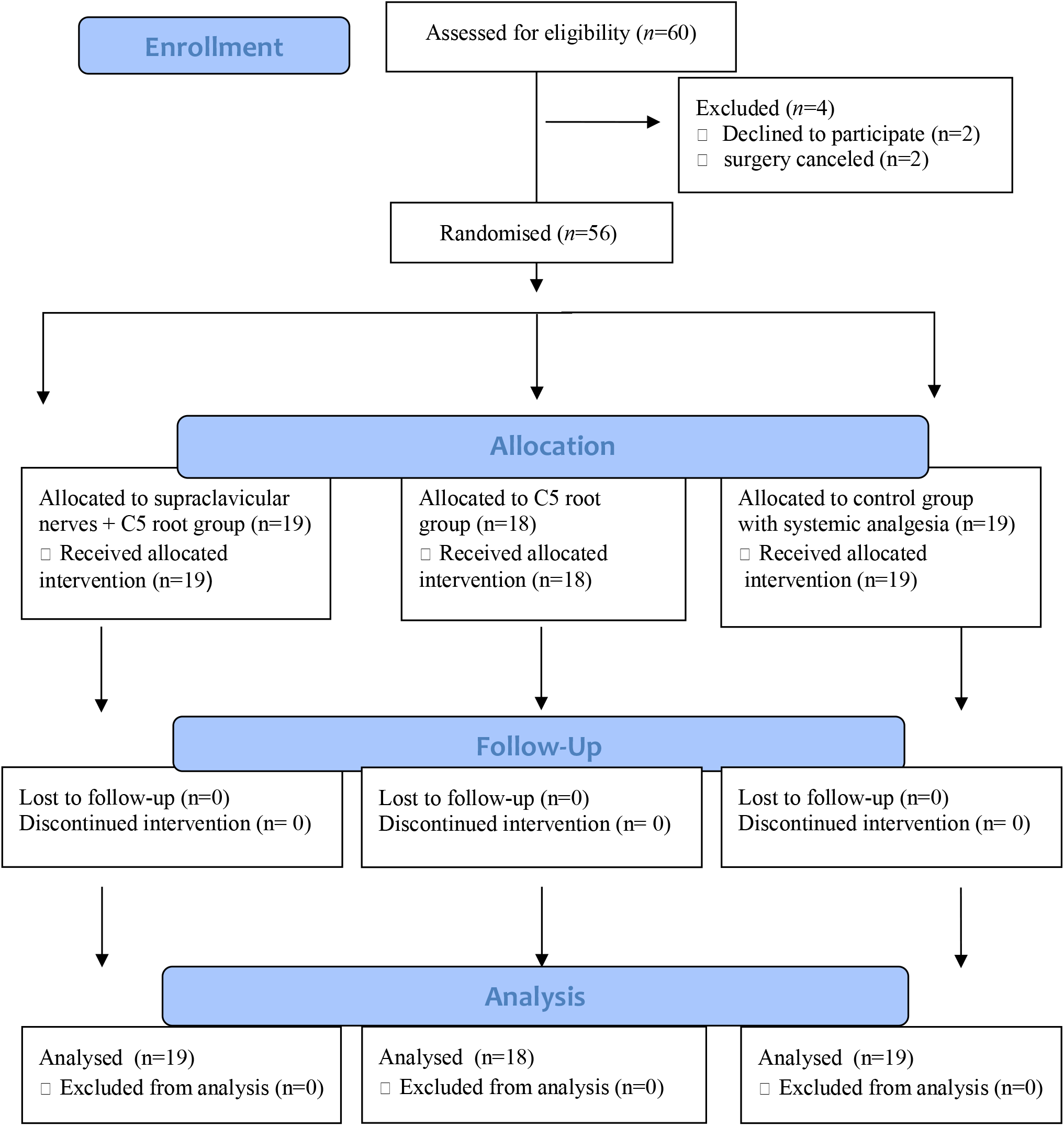
Flow chart according to CONSORT statement.

### Analgesic quality and opioid consumption

Patients who received a combined regional anesthesia of the supraclavicular nerves and the C5 nerve root reported no pain during the first postoperative hour (t0 to t1, NRS=0; Table 2). In contrast, patients in the control group experienced significantly higher pain levels at time points t0, t0.5, and t1 (*P*<0.001). The mean NRS score in the control group peaked at t0 with 4.5 ± 2.2, decreasing slightly to 3.2 ± 1.8 by t1. At t24, the NRS score in the NSCL+C5 group was 1.9 ± 0.9, marginally higher than in the control group (1.8 ± 1.0), although this difference was not statistically significant.

**Table 2.** Pain scores and piritramide consumption.

|  | NSCL+C5<br>(n=19) | C5<br>(n=18) | control<br>(n=19) | P-value<br>(NSCL+C5<br>vs control) | P-value<br>(C5 vs.<br>control) | P-value<br>(NSCL+C5<br>vs. C5) |
| --- | --- | --- | --- | --- | --- | --- |
| NRS t0 | 0 | 1.9 ± 1.4; (1.5) | 4.5 ± 2.2; (4) | <0.001 | <0.001 | 0.001 |
| NRS t0.5 | 0 | 2.9 ± 1.6; (3) | 4.4 ± 1.4; (4) | <0.001 | 0.002 | <0.001 |
| NRS t1 | 0 | 2.2 ± 1.2; (2) | 3.2 ± 1.1; (3) | <0.001 | 0.008 | <0.001 |
| NRS t24 | 1.9 ± 0.9; (2) | 2 ± 1; (2) | 1.8 ± 1; (2) | 1.000 | 1.000 | 1.000 |
| piritramide<br>(t0-t24) | 0 mg | 4.4 mg ± 5,3 | 12.6 mg ± 5.3 | <0.001 | <0.001 | 0.010 |

*Values are expressed as mean ± SD; (median), NRS=numeric rating scale; t0=first assessment postsurgery, t0.5=30 minutes postsurgery, t1=60 minutes postsurgery, t24=24 hours postsurgery, NSCL+C5=combined supraclavicular nerves and C5 root blockade; C5=C5 root blockade, control=systemic analgesia group*

The C5 group also reported low pain scores during the first postoperative hour. The highest pain intensity occurred at t0.5 with a mean of 2.9 ± 1.6, yet this remained significantly lower than that in the control group. The most notable difference between the C5 and control group was observed at t0 (1.9 vs. 4.5; *P*<0.001). At t24, the NRS score in the C5 group was 2.0 ± 1.0, slightly higher than that in the control group (1.8 ± 1), with no significant difference.

Comparison between the two intervention groups revealed significant differences at t0, t0.5, and t1 (t0: *P*=0.001; t0.5 and t1: *P*<0.001), with pain levels rising notably in the C5 group after the initial postoperative period. At t24, the NRS in the NSCL+C5 group remained lower at 1.9 ± 0.9 compared to 2.0 ± 1.0 in the C5 group, though the difference was not statistically significant.

None of the patients in the NSCL+C5 group required additional piritramide treatment throughout the study period (t0 to t24, Table 2). In contrast, the control group had a significantly higher mean cumulative piritramide consumption of 12.6 mg ± 5.3 (*P*<0.001). The C5 group required an average of 4.4 mg ± 5.3, which was also significantly lower than that in the control group (*P*<0.001). However, compared to the NSCL+C5 group, opioid consumption in the C5 group was significantly higher (*P*=0.010).

### Diaphragmatic excursion and oxygen saturation

The NSCL+C5 group exhibited a marked reduction in ipsilateral diaphragmatic excursion from t0 to t1 (Table 3). At t0, movement averaged 48.8% ± 35.7 of the baseline value, which was significantly lower than that in the control group (83.0% ± 11.0; P=0.001). Although slight improvements were observed at t0.5 (54.3% ± 37.5) and t1 (58.1% ± 38.6), values remained significantly reduced compared with the control group (t0.5: 80.3% ± 10.0; t1: 82.1% ± 9.4). By t24, diaphragmatic function was normalized in both groups. The C5 group also showed reduced diaphragmatic excursion during the first postoperative hour, with the lowest value recorded at t0 (63.6% ± 25.8 vs. 83.0% ± 11.0 in the control group), although this difference was not statistically significant (*P*=0.087). Values improved progressively at t0.5 (69.9% ± 20.6) and t1 (74.2% ± 15.8), remaining below those of the control group but without statistical significance. At t24, diaphragmatic movement had returned to baseline (95.4% ± 6.8). In a direct comparison, the NSCL+C5 group demonstrated a more pronounced—though not statistically significant—reduction in early postoperative diaphragmatic excursion.

**Table 3.** Diaphragmatic excursion and oxygen saturation.

|  | NSCL+C5<br>(n=19) | C5<br>(n=18) | control<br>(n=19) | P-value<br>(NSCL+C5<br>vs. control) | P-value<br>(C5 vs.<br>control) | P-value<br>(NSCL+C5<br>vs. C5) |
| --- | --- | --- | --- | --- | --- | --- |
| DE t0 | 48.8 ± 35,7 | 63.6 (± 25.8) | 83 ± 11 | 0.001 | 0.087 | 0.273 |
| DE t0,5 | 54.3 ± 37.5 | 69.9 ± 20.6 | 80.3 ± 10 | 0.008 | 0.654 | 0.205 |
| DE t1 | 58.1 ± 38.6 | 74.2 ± 15.8 | 82.1 ± 9.4 | 0.013 | 1 | 0.163 |
| DE t24 | 100.2 ± 10.5 | 95.4 ± 6.8 | 99.1 ± 5.1 | 1 | 0.494 | 0.204 |
| SpO<sub>2</sub> t0 | 98.4 ± 1.4 | 98 ± 1.9 | 96.9 ± 1.8 | 0.048 | 0.245 | 1 |
| SpO<sub>2</sub> t0.5 | 99.1 ± 1.1 | 98.2 ± 1.7 | 97.7 ± 1.9 | 0.019 | 0.940 | 0.240 |
| SpO<sub>2</sub> t1 | 99.6 ± 0.8 | 98.7 ± 1.6 | 97.3 ± 1.9 | <0.001 | 0.017 | 0.196 |
| SpO<sub>2</sub> t24 | 100.1 ± 1 | 99.3 ± 1 | 99.4 ± 1 | 0.136 | 1 | 0.080 |

*Values are expressed as mean ± SD in percent (%) of preoperative baseline, DE=diaphragmatic excursion, t0=first assessment postsurgery, t0.5=30 minutes postsurgery, t1=60 minutes postsurgery, t24=24 hours postsurgery, NSCL+C5=combined supraclavicular nerves and C5 root blockade; C5=C5 root blockade, control=systemic analgesia group*

Patients in the NSCL+C5 group demonstrated significantly higher oxygen saturation during the first postoperative hour (t0–t1) compared with the control group, with the most pronounced difference noted at t1 (99.6% ± 0.8 vs. 97.3% ± 1.9, Table 3). While both groups experienced a minor decline from baseline, saturation in the NSCL+C5 group improved steadily, whereas the control group exhibited a transient dip at t1. At t24, oxygen saturation returned to baseline in both groups, with no statistically significant difference (NSCL+C5: 100.1% ± 1.0 vs. control: 99.4% ± 1.0). The C5 group also showed higher oxygen saturation values than the control group from t0 to t1, however, only the value at t1 reached statistical significance (98.7% ± 1.6 vs. 97.3% ± 1.9; *P*=0.017). At t24, both groups returned to near-baseline saturation levels (C5: 99.3% ± 1.0; control: 99.4% ± 1.0). Although the NSCL+C5 group consistently demonstrated higher saturation values than the C5 group, these differences were not statistically significant.

### Harms and unintended effects

The assessment of adverse events focused primarily on the occurrence of subjective dyspnea at rest as a potential indicator of pulmonary impairment. None of the patients reported dyspnea at rest at any time point, either pre-operatively or postoperatively (t0 to t24). In addition to the aforementioned cases of phrenic nerve palsy, no additional complications or unintended effects were observed.

## Discussion

### Analgesic quality and opioid consumption

This study demonstrated that a combined regional anesthesia targeting the supraclavicular nerves and C5 nerve root can provide highly effective analgesia for clavicle fractures and acromioclavicular joint dislocations. All patients in the NSCL+C5 group remained entirely pain-free during the immediate postoperative period (NRS=0). Given the ongoing lack of standardized analgesic protocols for clavicle injuries, this finding is clinically significant and has led to the implementation of this approach as a standard practice within our institution.

Postoperative pain is typically high in trauma and orthopedic procedures, with many patients requiring substantial analgesia during the first 24 hours.^12^ This pattern was evident in the control group, where patients reported median NRS scores of 4 at both t0 and t0.5. Adequate pain relief was achieved only after substantial administration of piritramide. Patients in the C5 group experienced more pain than those in the NSCL+C5 group, although NRS scores never exceeded three, indicating only mild discomfort. These findings suggest that a selective C5 block alone provides superior analgesia compared with systemic opioids; however, complete pain control was only achieved when the supraclavicular nerves were additionally blocked.

While several authors have previously highlighted the importance of blocking both cervical and brachial plexus components, there is currently no consensus regarding the optimal regional anesthesia technique for clavicle surgery.^3,4^ Our findings support the high analgesic efficacy of a combined, selective supraclavicular nerve and C5 root block. This aligns with previous reports, such as that by Valdés-Vilches et al., who demonstrated effective analgesia using a supraclavicular nerve block, although combined with a supraclavicular brachial plexus approach rather than a selective C5 root block.^13^ Similarly, case reports by Kline and Shanthanna described successful pain control using combinations of superficial cervical plexus and selective C5 root blocks.^14,15^ With the advancement of ultrasound-guided techniques, newer approaches such as the clavipectoral fascial plane block have also gained attention as promising alternatives for postoperative analgesia.^16,17^ Beyond postoperative analgesia, regional anesthesia has been employed not only for postoperative pain control but also as a sole anesthetic technique for clavicle surgery.^18,19^ For instance, Balaban et al. retrospectively reported 12 successful procedures using combined cervical and interscalene blocks.^18^ However, as our study focused exclusively on postoperative outcomes, intraoperative efficacy could not be evaluated. A prospective, randomized trial examining the use of selective blockade of the supraclavicular nerves and the C5 root—without general anesthesia—would be of particular interest, especially in the context of patient comfort and enhanced recovery concepts.

Importantly, none of the patients in the NSCL+C5 group required supplemental opioids throughout the 24-hour postoperative period, in contrast to average piritramide doses of 4.4 mg in the C5 and 12.6 mg in the control group. In the context of the global opioid crisis, this regional anesthesia technique may represent a significant advancement in reducing perioperative opioid use and enhancing patient safety.^20,21^

### Diaphragmatic excursion and oxygen saturation

Patients in the NSCL+C5 group showed significantly reduced diaphragmatic excursion on the operated side during the first postoperative hour. This finding was consistent with the anatomical proximity of the phrenic nerve (C3–C5) to the injection site. However, reduced movement may not solely result from nerve involvement, general anesthesia itself can transiently affect diaphragmatic function.^22^ Although subsequent studies have demonstrated that reducing the volume of local anesthetic and modifying the injection technique can significantly lower the risk, a transient phrenic nerve palsy cannot be entirely ruled out when using these blockade techniques.^23,24^ Notably, Wiesmann et al. found lower rates of phrenic palsy with supraclavicular blocks compared with interscalene blocks.^24^ Similarly, Shanthanna described a case of effective analgesia using combined selective C5 and cervical plexus block, without diaphragmatic impairment.^15^ Fascial plane blocks, such as the clavipectoral block, may also reduce phrenic nerve involvement while maintaining adequate analgesia.^16^

Despite reduced diaphragmatic movement, no oxygenation impairment was observed. In fact, the NSCL+C5 group exhibited higher postoperative oxygen saturation values, likely due to superior pain control and reduced opioid consumption. Unilateral diaphragm paresis is typically asymptomatic in healthy patients, as compensation via the contralateral diaphragm and accessory muscles maintains adequate ventilation.^9^ However, our findings may not apply to patients with pre-existing pulmonary disease or obesity, where compensatory capacity may be limited.

## Data Availability

All data produced in the present study are available upon reasonable request to the authors

## Limitations

In addition to the limitations already discussed, the non-blinded randomization of the study design may have introduced systematic bias. Furthermore, generalizability is limited by the single-center design and the absence of patients with severe pulmonary comorbidities.

## Conclusion

Based on these results, we conclude that combined selective regional anesthesia of the supraclavicular nerves and the C5 root may be an effective method for preventing postoperative pain and significantly reducing opioid consumption in clavicle surgery. Although there was an increased incidence of phrenic nerve palsy, leading to a reduction in diaphragmatic movement in the NSCL+C5 group, no adverse effects on oxygen saturation or clinically relevant signs of desaturation were observed. Future studies are needed to investigate the efficacy of this regional anesthesia technique without additional general anesthesia.

## References

1. Kihlström C, Möller M, Lönn K, Wolf O. Clavicle fractures: epidemiology, classification and treatment of 2 422 fractures in the Swedish Fracture Register; an observational study. BMC Musculoskelet Disord. 2017;18(1):82. doi: 10.1186/s12891-017-1444-1

2. Epstein D, Day M, Rokito A. Current concepts in the surgical management of acromioclavicular joint injuries. Bull NYU Hosp Jt Dis. 2012;70(1):11–24.

3. Ryan DJ, Iofin N, Furgiuele D, Johnson J, Egol K. Regional anesthesia for clavicle fracture surgery is safe and effective. J Shoulder Elb Surg. 2021;30(7):e356–60. doi: 10.1016/j.jse.2020.10.009

4. Banerjee S, Acharya R, Sriramka B. Ultrasound-guided interscalene brachial plexus block with superficial cervical plexus block compared with general anesthesia in patients undergoing clavicular surgery: a comparative analysis. Anesth Essays Res. 2019;13(1):149–54. doi: 10.4103/aer.AER_185_18

5. Döffert J, Steinfeldt T. Anästhesie in der Orthopädie/Unfallchirurgie – Regionalanästhesie bei Verletzungen der oberen Extremität. Anasthesiol Intensivmed Notfallmed Schmerzther. 2015;50(4):270–8. doi: 10.1055/s-0040-100152

6. Tran DQH, Tiyaprasertkul W, Gonzalez AP. Analgesia for clavicular fracture and surgery: a call for evidence. Reg Anesth Pain Med. 2013;38(6):539–43. doi: 10.1097/AAP.0000000000000012

7. Albrecht E, Chin KJ. Advances in regional anesthesia and acute pain management: a narrative review. Anaesthesia. 2020;75(S1):e101–10. doi: 10.1111/anae.14868

8. American Society of Anesthesiologists Task Force on Acute Pain Management. Practice guidelines for acute pain management in the perioperative setting: an updated report. Anesthesiology. 2012;116(2):248–73. doi:10.1097/ALN.0b013e31823c1030

9. El-Boghdadly K, Chin KJ, Chan VWS. Phrenic nerve palsy and regional anesthesia for shoulder surgery: anatomical, physiologic, and clinical considerations. Anesthesiology. 2017;127(1):173–91. doi: 10.1097/ALN.0000000000001668

10. Hogan QH. Phrenic nerve function after interscalene block revisited: now, the long view. Anesthesiology. 2013;119(2):250–2. doi: 10.1097/ALN.0b013e31829c2f3a

11. Rhyner P, Kirkham K, Hirotsu C, Farron A, Albrecht E. A randomised controlled trial of shoulder block vs interscalene brachial plexus block for ventilatory function after shoulder arthroscopy. Anaesthesia. 2020;75(4):493–498.e1. doi:10.1111/anae.14957

12. Gerbershagen HJ, Aduckathil S, van Wijck AJM, Peelen LM, Kalkman CJ, Meissner W. Pain intensity on the first day after surgery: a prospective cohort study comparing 179 surgical procedures. Anesthesiology. 2013;118(4):934–44. doi: 10.1097/ALN.0b013e31828866b3

13. Valdés-Vilches LF, Sánchez-del Águila MJ. Anesthesia for clavicular fracture: selective supraclavicular nerve block is the key. Reg Anesth Pain Med. 2014;39(3):258–9. doi:10.1097/AAP.0000000000000057

14. Kline JP. Ultrasound-guided placement of combined superficial cervical plexus and selective C5 nerve root catheters: a novel approach to treating distal clavicle surgical pain. AANA J. 2013;81(1):19–22.

15. Shanthanna H. Ultrasound-guided selective cervical nerve root block and superficial cervical plexus block for surgeries on the clavicle. Indian J Anaesth. 2014;58(3):327–9. doi: 10.4103/0019-5049.135050

16. Zhuo Q, Zheng Y, Hu Z, Xiong J, Wu Y, Zheng Y, et al. Ultrasound-guided clavipectoral fascial plane block with intermediate cervical plexus block for midshaft clavicular surgery: a prospective randomized controlled trial. Anesth Analg. 2022;135(3):633–40. doi: 10.1213/ANE.0000000000005911

17. Pereira CS, Ferros C, Dinis I, Pereira D, Miguel D, Vico M. Clavipectoral fascial plane block for clavicle fracture surgery: a case report. J Perioper Pract. 2024;34(12):375–7. doi: 10.1177/17504589241264408

18. Balaban O, Dülgeroğlu TC, Aydın T. Ultrasound-guided combined interscalene-cervical plexus block for surgical anesthesia in clavicular fractures: a retrospective observational study. Anesthesiol Res Pract. 2018;2018:7842128. doi:10.1155/2018/7842128

19. Baran O, Kir B, Ateº I, Şahin A, Üztürk A. Combined supraclavicular and superficial cervical plexus block for clavicle surgery. Korean J Anesthesiol. 2020;73(1):67–70. doi: 10.4097/kja.d.18.00296

20. Kumar K, Kirksey MA, Duong S, Wu CL. A review of opioid-sparing modalities in perioperative pain management: methods to decrease opioid use postoperatively. Anesth Analg. 2017;125(5):1749–60. doi: 10.1213/ANE.0000000000002497

21. Trasolini NA, McKnight BM, Dorr LD. The opioid crisis and the orthopedic surgeon. J Arthroplasty. 2018;33(11):3379–82.e1. doi:10.1016/j.arth.2018.07.002

22. Ricoy J, Rodríguez-Núñez N, Álvarez-Dobaño JM, Toubes ME, Riveiro V, Valdés L. Diaphragmatic dysfunction. Pulmonology. 2019;25(4):223–35. doi: 10.1016/j.pulmoe.2018.10.008

23. Lee JH, Cho SH, Kim SH, Chae WS, Jin HC, Lee JS, et al. Ropivacaine for ultrasound-guided interscalene block: 5 mL provides similar analgesia but less phrenic nerve paralysis than 10 mL. Can J Anaesth. 2011;58(11):1001–6. doi: 10.1007/s12630-011-9568-5

24. Wiesmann T, Feldmann C, Müller HH, Nentwig L, Beermann A, El-Zayat BF, et al. Phrenic palsy and analgesic quality of continuous supraclavicular vs interscalene plexus blocks after shoulder surgery. Acta Anaesthesiol Scand. 2016;60(8):1142–51. doi: 10.1111/aas.12732

